# Complete pharmacogenomic profile from exome sequencing

**DOI:** 10.64898/2026.01.13.26343772

**Authors:** Ilias Bensouna, Aleksandra Grujic, Fanny Ponce, Nicolas Jauniaux, Tanja Scheikl, Nicolas Picard, Boris Chaumette, Karl-Dietrich Hatz, Xavier Vanhoye, Laurent Mesnard, Laure Raymond

**Affiliations:** Soins Intensifs Néphrologiques et Rein Aigu (SINRA), Nephrology Department, Tenon Hospital, Assistance Publique – Hôpitaux de Paris, Paris, France; Inserm UMR_S1155, Paris, France; Eurofins Biomnis Laboratory, Lyon, France; Université Paris Cité, Institute of Psychiatry and Neuroscience of Paris (INSERM U1266); INTLAB AG, 8707 Uetikon am See, Switzerland; Department of Pharmacology, Toxicology and Pharmacovigilance, CHU de Limoges, Limoges, France; Pharmacology & Transplantation, INSERM U1248, Université de Limoges, Limoges, France; Department of Psychiatry, McGill University, Montreal, Canada; Institut Pasteur (CNRS UMR3571), GHU Paris Psychiatrie et Neurosciences, Paris, France; Centre Maladie Rare MAHREA, Tenon Hospital, Assistance Publique – Hôpitaux de Paris, Paris, France; Medicine Faculty, Paris Sorbonne University, Paris, France; European Rare Kidney Disease Reference Network (ERKNet), Heidelberg, Germany

**Keywords:** pharmacogenetics, whole exome sequencing, HLA, *CYP2D6*

## Abstract

Exome sequencing (ES) is a cornerstone of clinical genetic diagnosis, yet its application in pharmacogenomics remains limited. While some pharmacogenetic variants are detectable by ES, clinically relevant loci such as *CYP2D6*, *UGT1A1*, and HLA remain challenging. We present a robust, comprehensive method to derive a complete pharmacogenomic profile directly from standard ES data. Our method addresses primary limitations of ES for pharmacogenomics, including low coverage and structural complexity at critical loci. We analyzed 66 samples from diverse sources, targeting 217 variants across a panel of 23 pharmacogenes. The method was validated by comparing its results with reference samples from the Genetic Testing Reference Material Coordination Program, as well as with the Veridose Core+CNV assay® (Agena) and the Personal Medicine Profile assay® (GeneTelligence). HLA typing performance was assessed and confirmed through comparison with both the Immucor LIFECODES HLA-SSO kit and a clinical transplantation-grade HLA assay. This validation demonstrates that ES can provide a comprehensive pharmacogenomic profile in a single, streamlined workflow, facilitating seamless integration of pharmacogenomics into precision medicine.

## Introduction

Pharmacogenomics is a field of genomic that aims to optimize and individualize drug treatment based on DNA analysis to increase drug efficacy and prevent adverse effects. A recent study from a large cohort published by the Ubiquitous Pharmacogenomic Consortium (U-PGx) demonstrates the interest of pre-emptive pharmacogenetic panel testing, with a 30% reduction rate of clinically detectable side-effects^1^.

For the time being, pharmacogenetic analysis relies on two main strategies. On the one hand, highly targeted approaches are used, such as real-time PCR focusing on selected variants. On the other hand, more comprehensive multi-locus approaches are employed to produce larger pharmacogenetic profile. The latter relies on the use of specific techniques, such as the DNA microarray, mass spectrometry coupled with allele-specific PCR, or second generation sequencing panels.^2,3^ A method that would enable simultaneous pharmacogenetic profiling and molecular diagnosis using a single technology, while minimizing biases such as the allele drop-out commonly associated with PCR, would be of particular interest.

To date, the use of exome sequencing (ES) to obtain pharmacogenetic profiles has never been fully validated, particularly for certain pharmacogenes. An initial study of 1928 variants from 62 samples studied in ES resulted in 14% false negatives, mainly due to insufficient sequencing depth^4^. While it has been possible to identify part of the variants in pharmacogenes, there are difficulties associated with certain loci. A recent study of 19 pharmacogenes concluded that ES data was unusable for 7 of them^5^. Relevant pharmacogene variants are located in intronic regions and are therefore not typically captured by ES^6^. ES has traditionally been considered unsuitable for analyzing the *CYP2D6* gene – involved in the metabolism of 25% of drugs – due to its high sequence similarity with pseudogenes and the presence of numerous CNVs or hybrid genes^7–9^. A pre-amplification using long-range PCR is done to improve sequencing of this region but it does not allow the detection of hybrid genes^10^. Moreover, difficulties have also been encountered for the sequencing of TA repeats in the *UGT1A1* promoter, as well as for the detection of certain variants located in *CYP3A5* and *CYP2C19*^11^. In addition, there have been few studies to date on HLA profiling by ES, but this method is beginning to be tested, particularly in determining the association of HLA profiles with autoimmune diseases^12^.

Beyond providing a molecular diagnosis, ES offers the opportunity to extract additional information yet underutilized, particularly on the pharmacogenetic profile. With the advent of second-generation sequencing, thousands of patients are now benefiting from ES as part of routine diagnostic practice. It would therefore be cost-effective to recycle this data instead of performing a complementary technique, with the possibility of answering both questions in a single step. The variants of interest in pharmacogenes are located either in exonic regions or in adjacent intronic regions, except for a few variants^13^. On top of giving information for exonic regions, ES also provides the sequence of those border intronic regions, and can theoretically cover most of the variants of interest. Our aim is therefore to validate the use of this test to establish a complete pharmacogenomic profile, providing additional information from a single test.

## Material and methods

### Variants and genes of interest for pharmacogenetic analysis

The variants and genes of interest for pharmacogenetic analysis were selected with the help of the guidelines from the Clinical Pharmacogenetics Implementation Consortium (CPIC) and the Dutch Pharmacogenetics Working Group (DPWG), as well as relevant drug label information. Our panel was designed based on the PGx Working Group’s recommendations for a minimum set (Tier 1) and an extended set (Tier 2).^14–20^ A total of 217 variants spanning 23 pharmacogenes were selected for their frequencies and their effect on the variability of drug response of at least one therapeutic drug. The potential impact of these variants on protein structure, expression and/or function has already been studied. A summary of the tested alleles and genes is presented in **Table 1**. The panel serves as a proof-of-concept for the analysis of exome sequencing data.

**Table 1:** Panel of pharmacogenes with their alleles selected for pharmacogenomic analysis from ES data.

| Gene | Tested alleles |
| --- | --- |
| **CYP2B6** | *2, *4, *5, *6A, *7, *8, *9, *12, *13, *16, *18, *19, *20, *22, *26, *28, *34, *35, *38 |
| **CYP2C9** | *2, *3, *4, *5, *6, *8, *11, *12, *13, *15, *25, *27 |
| **CYP2C19** | *2, *3, *4A, *4B, *5, *6, *7, *8, *9, *10, *16, *17, *19, *22, *24, *25, *26, *35 |
| **CYP2D6** | *2, *3, *4.001, *4.009, *4.010, *4.012, *6, *6.003, *7, *8, *9, *10, *11, *12, *14.001, *15, *17, *18, *19, *20, *21, *29, *31, *32, *34, *36, *38, *39, *40, *41, *42, *44, *47, *49, *51, *52, *56.001, *56.002, *57, *59, *62, *68, *69, *81, *91, *92, *96, *100, *101, *114,<br>CNV: *5, x2, x3<br>Hybrid genes: *4.013, *13, *36, *61, *63, *68, *83 |
| **CYP3A5** | *3, *3.010, *6, *7 |
| **IFNL3** | rs12979860 C>T |
| **POR** | *28 |
| **TPMT** | *2, *3A, *3B, *3C, *4 |
| **UGT1A1** | *6, *7, *27, *28, *28c, *29, *36, *37 |
| **ABCG2** | 421A |
| **BCHE** | 382T, 467G, 895T, 1004C, 1354T, 1584A, A, AK, F1, F2, F2Sc, F Japan, J, JK, K, Sc, Silent7 |
| **CACNA1S** | 520T, 3257A |
| **CYP3A4** | *2, *17, *22 |
| **CYP4F2** | *3 |
| **DPYD** | *2A, *3, *7, *8, *10, *12, *13, 557G, 2846T, HapB3 |
| **G6PD** | A, A- 202A_376G, A- 680T_376G, A- 968C_376G, Asahi, Aures, Belem, Canton, Chatham, Chinese-5, Coimbra Shunde, Cosenza, Gaohe, Guadalajara, Hechi, Ilesha, Kaiping, Kalyan-Kerala, Kamiube, Kamogawa, Lagosanto, Liuzhou, Ludhiana, Mahidol, Malaga, Mediterranean, Mexico City, Montalbano, Montpellier, Nanning, Orissa, Palestrina, Quing Yan, Radlowo, Santa Maria, Santiago, Seattle, Sibari, Sierra Leone, Sinnai, Sumare, Taipei, Ube Konan, Union, Vanua Lava, Viangchan |
| **MT-RNR1** | 1095C, 1494T, 1555G |
| **NUDT15** | *3 |
| **RYR1** | 103C, 130T, 487T, 488T, 742A/C, 982T, 1021A/C, 1201T, 1209G, 1565C, 1589A, 1597T, 1598A, 1654T, 1840T, 1841T, 6487T, 6488A, 6502A, 6617G/T, 7007A, 7039delGAG, 7048A, |
|  | 7063T, 7124C, 7282A, 7300A, 7304A, 7354T, 7360T, 7361A, 7372T, 7373A, 7522A/T,<br>7523A, 9310A, 11969T, 14387G, 14477T, 14497T, 14512G, 14545A, 14582A, 14693C |
| *SLCO1B1* | *5 |
| *VKORC1* | -1639G>A |

### Sample collection

A total of 66 samples were included to validate the use of ES in pharmacogenomic profile. 6 reference samples, specifically chosen for their challenging characteristics, were purchased at Coriell Institute (NA12878, HG00421, HG01190, NA07439, NA10860 and NA18545). For those samples, trust sets are available for *CYP2D6* star allele assignment, HLA typing, and genome sequencing (GS)^21–23^. These 6 reference samples are samples from cell lines, extensively characterized for up to 28 pharmacogenes by the Genetic Testing Reference Material Coordination Program (GeT-RM)^24^. 33 samples were sent to the Agena laboratory to perform the Veridose Core+CNV assay®, based on PCR amplification and LC-MS detection^25^. 7 clinical samples and 1 reference DNA (NA12878) were sent to GeneTelligence to perform the Personal Medicine Profile assay (GeneTelligence, based on Illumina® Infinium™ Global Diversity Array platform (PMP)). 21 other samples were used to evaluate the performance for HLA allele detection: 14 samples were analyzed using the Immucor LIFECODES® HLA-SSO Typing kit, which uses sequence specific oligonucleotides to identify which HLA alleles are present in a PCR-amplified sample. 7 other samples had an HLA result for transplantation purposes from the Immunology and Histocompatibility laboratory of Saint-Louis Hospital, based on Next Generation Sequencing method. Flowchart of the proof-of-concept is described in **Figure 1**.

**Figure 1:**
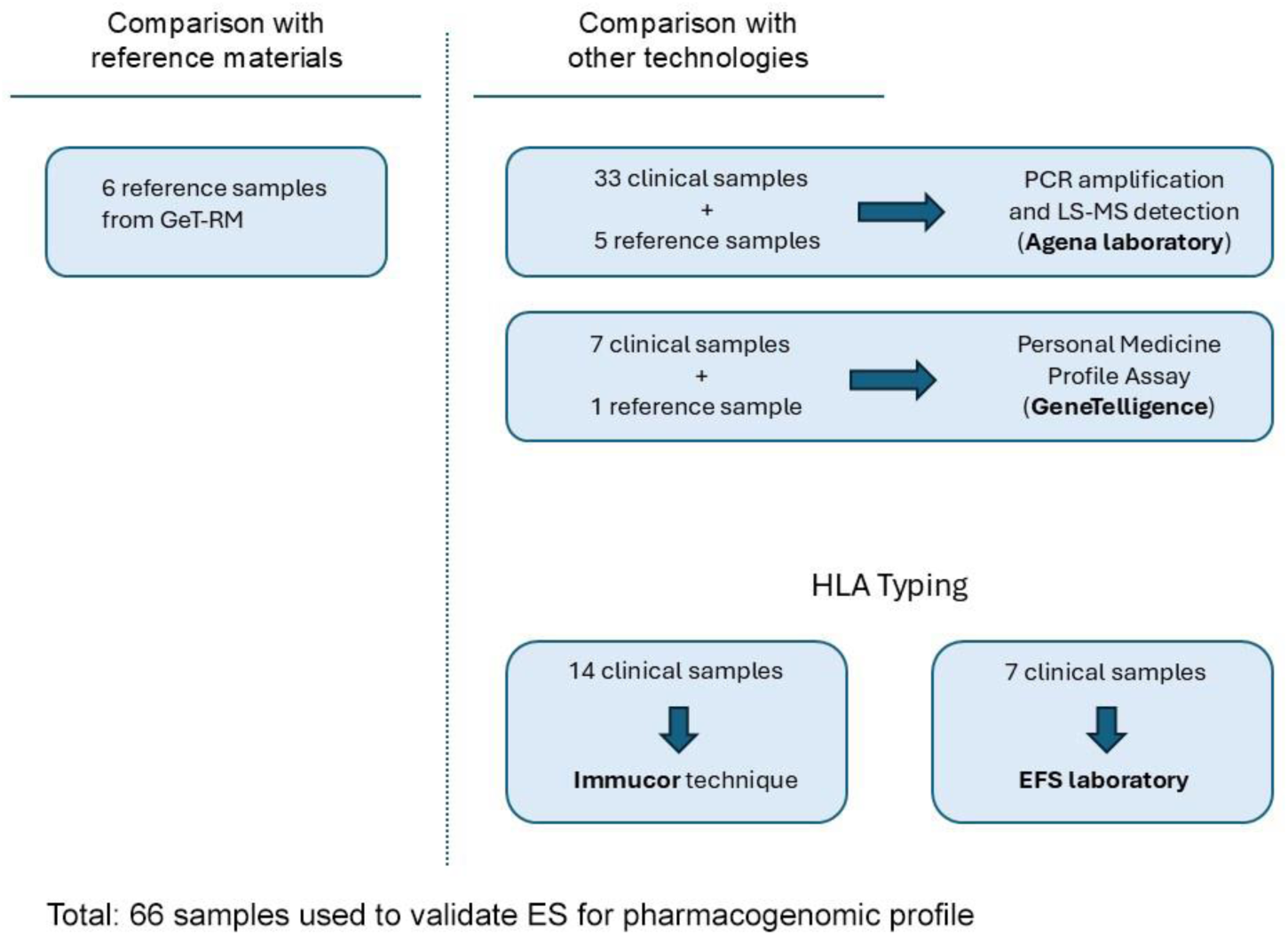
Flowchart of samples used to validate the use of ES for determination of pharmacogenomic profile. One sample was used both for comparison with reference samples and for comparison with LC-MS detection. Another sample was used both for comparison with GeneTelligence and for comparison with Immucor. LC-MS detection was performed by Agena laboratory, GeneTelligence laboratory performed the Personal Medicine Profile assay for 1 reference sample from GeT-RM group + 7 other samples. *GeT-RM = Genetic Testing Reference Material Coordination Program; EFS: « établissement français du sang »; LC-MS = Liquid Chromatography coupled to tandem Mass Spectrometry*

Some differences inherent to reference methods were expected: alleles *2, *9 and *36 of *CYP2D6* are not tested by PMP assay, nor *4, *5, *6 and *9 of *DPYD*. Moreover, *80 of *UGT1A1* gene is tested with PMP assay (instead of *28), which is a proxy for *28 (detected by in-house pipeline).^26^

### ES sequencing and standard bioinformatics

The analysis is carried out on peripheral blood collected in an EDTA blood tube or DNA. If needed, genomic DNA was isolated from EDTA blood using the DNA Minikit from QIAGEN SA. From 50 ng of enzymatically fragmented DNA, indexed libraries were prepared and hybridized with biotinylated probes of Twist Comprehensive Exome before May 2023, and Twist Exome 2.0 plus Comprehensive Exome Spike-in after^27^. The samples were prepared following the manufacturer recommendations. The pools of libraries were sequenced on the Illumina NovaSeq 6000 sequencer in paired end (2x150 bp). The raw data were converted to fastq format using BCL Convert software. The sequences are analyzed following the best practices recommended by the GATK Broad Institute, with an in-house pipeline based on BWAmem for alignment against GRCh38 reference genome, HaplotypeCaller for small variant calling, and GermlineCNVCaller for CNV calling^28,29^. Sequenced reads were visualized using IGV.

### Bioinformatic tools for pharmacogenomics analysis

In-house scripts were developed to collect relevant information from standard ES workflow and dedicated pharmacogenetic tools: single nucleotides variations (SNVs) and indels relevant to pharmacogenetics, determination of the number of TA repeats in the *UGT1A1* promoter, detection of *CYP2D6*::*CYP2D7* or *CYP2D7*::*CYP2D6* rearrangements. Structural variants are detected using read depth information. Control depth is calculated within a large cohort of 1246 samples that are used as control. For HLA-A and B typing, the tool xHLA was added to the pipeline. This tool is based on an algorithm that performs iterative refinement of protein-level alignments using precomputed multiple sequence alignments to achieve 99-100% accurate 4-digit HLA typing from short-read sequencing data^30^. Those in-house scripts provide for each sample of the batch the clinical and demographical information (extracted from our LIMS), the genotype of every SNV of interest (with depth above under 10X) and a CNV report enabling the detection of the presence of CNV in *CYP2D6* or *CYP2D6*/*CYP2D7* rearrangement. All this information is compiled in a csv file. This file comes with a BAM and an html files, allowing visual checking for suspicious variants; providing IGV view for SNV and bedgraph files for CNV^31,32^. From a CNV signal transcribed in the form of a karyoplot pattern, we can visually extrapolate the presence of a hybrid gene (**Figure 2**). Also, the *4 allele in *CYP2D6* must be manually curated using IGV because of the noise induced by the adjacent pseudogene. Another output file contains information on HLA-A and HLA-B results. The csv files were sent to INTLAB AG (Switzerland) which makes genotype and phenotype interpretation, and generates a PGx report with therapeutic recommendations based on established international guidelines and drug label information.

**Figure 2:**
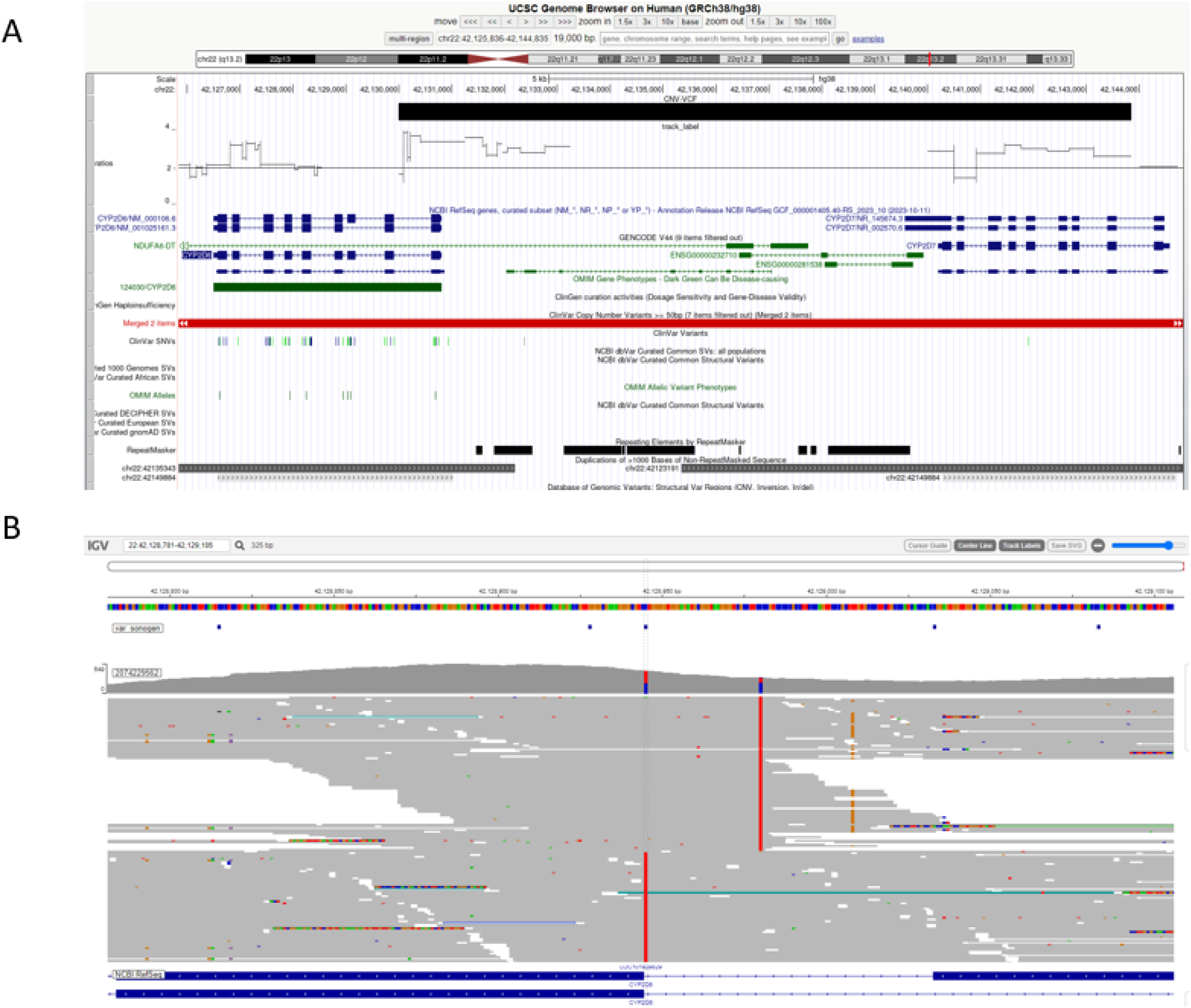
Visualization of the *CYP2D6* and *CYP2D7* region for a patient carrying a *CYP2D6*::*CYP2D7* hybrid gene. A) Bedgraph file loaded on USCS genome browser, showing a duplication occurring in exon 1 of *CYP2D6* and in exons 2 to 9 in *CYP2D7*, corresponding to the *CYP2D6*::*CYP2D7* hybrid *68. B) The sample has been called heterozygous for *CYP2D6* *4 (chr22:42128945C>T rs3892097). However, on IGV view from BAM file, wild reads are in fact pseudogene reads (with the two SNP on the right).

### Analytical performance

To measure the analytical sensitivity and specificity of this method, pharmacogenomic analysis results from ES data were compared with 3 consensus genotypes derived from the Get-RM group.

## Results

### Data quality

The in-house ES pipeline has been evaluated and validated in terms of accuracy, repeatability, and precision for routine molecular diagnosis according to COFRAC recommendations^33^. The sequenced data include exonic regions, the border part of intronic regions, upstream and downstream regions of human genome. The mean depth is 80X and 99.5% of bases have a depth above 10X, which is in line with the expected reference values. All 217 variants are covered by ES data with depth > 10X, since the Twist Exome 2.0 plus Comprehensive Exome Spike-in design also includes certain intronic regions of interest.

### Genotype validation with Whole Genome Sequencing for reference materials

When comparing the variant calling from ES data against variant calling from Whole genome Sequencing for the 6 reference (HG00421, HG01190, NA07439, NA10860, NA18545, NA12878), of the 217 genetic polymorphisms tested (SNPs and indels) for each sample (1314 variants in total), 1310 were identical^34^. The 4 discrepancies concern sample HG01190 and the *CYP2D6* gene. The BAM analysis of the genome sequencing shows that for these 4 positions, there is an error in the vcf file provided (**Table 2**), which is confirmed by the genotype published by the Get-RM for this same sample (*68+*4/*5). The concordance is therefore 100% (1314/1314).

**Table 2:** Summary of discrepancies between genome sequencing reference data and exome sequencing data, for sample HG01190. For the 4 discrepancies found in the vcf file, a visual study of the BAMs led to the conclusion that the error came from the vcf file supplied and that the sequence finally corresponded to the genotype published by Get-RM.

| RSID | CHROM :POS (HG38) | VCF GENOME | BAM GENOME | CORRECTED GENOTYPE | IN-HOUSE DATA | CONCLUSION |
| --- | --- | --- | --- | --- | --- | --- |
| RS16947 | chr22 :42127941 | G/A | chr22:42 127 941<br>Total count: 30<br>A : 0<br>C : 0<br>G : 30 (100%, 7+, 23- )<br>T : 0<br>N : 0 | G/G | G/G | Compliant |
| RS1065852 | chr22:42130692 | G/A | chr22:42 130 692<br>Total count: 52<br>A : 52 (100%, 25+, 27- )<br>C : 0<br>G : 0<br>T : 0<br>N : 0 | A/A | A/A | Compliant |
| RS2267447 | chr22:42128694 | T/C | chr22:42 128 694<br>Total count: 15<br>A : 0<br>C : 15 (100%, 10+, 5- )<br>G : 0<br>T : 0<br>N : 0 | C/C | C/C | Compliant |
| RS2837172<br>5 | chr22:42127803 | T/C | chr22:42 127 803<br>Total count: 35<br>A : 0<br>C : 35 (100%, 18+, 17- )<br>G : 0<br>T : 0<br>N : 0 | C/C | C/C | Compliant |

### Star allele assignment for reference materials

For 3 samples available in house (NA12878, HG01190, NA07439), extensive analysis was performed by GeT-RM to identify the truth star allele calling for 11 genes^24^. All the alleles of the studied pharmacogenes were consistent between the reference data and the locally sequenced data. **Table 3** details the comparison between the data obtained by ES and the Get-RM reference data.

**Table 3:**
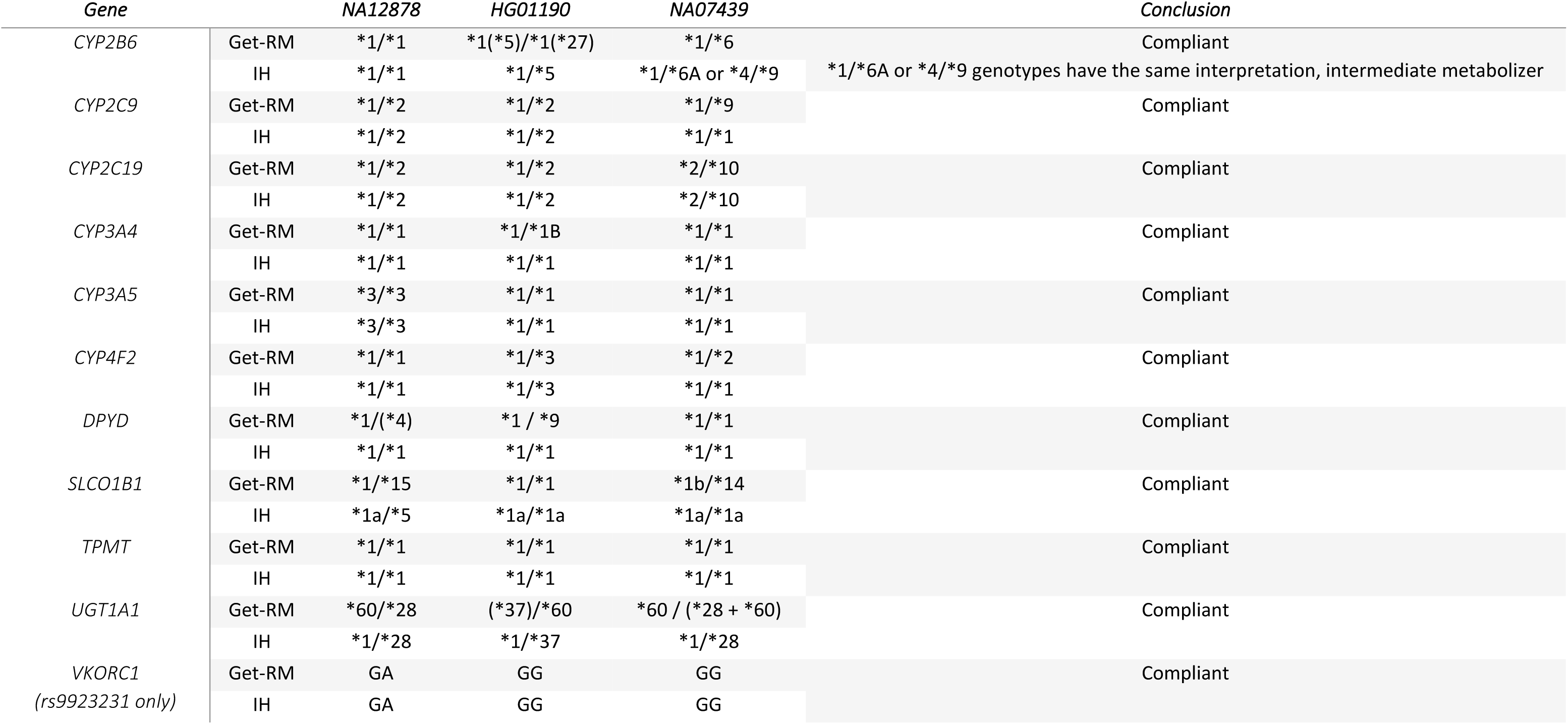
Star allele assignment for 3 reference samples (Get-RM) compared with IH sequencing. For *CYP2B6*: Alleles in parentheses indicate that these alleles have not been confirmed by a separate method (some alleles were included in only one assay because of differences in platform design). For sample HG01190, another study, based on WGS, lead to identification of *1/*5.^24^ For *CYP2C9* and *DPYD*, *9 leads to normal function and is not assessed by in-house pipeline. For *CYP3A4*, *1B is now wild type and named *1.001. For CYP4F2, the V433M (*3) variant is the only one considered for warfarin recommendations, *2 was not assessed by in-house pipeline For *SLCO1B1*, *1b corresponds to *37 in the new nomenclature (normal function), *5 and *15 share the same variant and are both nonfunctional. *15 is not assessed by in-house pipeline. *14 has increased function and is not assessed by in-house pipeline. For *UGT1A1*, *60 leads to normal function and is not assessed by in-house pipeline.

### Comparison with other technologies

For the 8 samples sent to GeneTelligence to perform the Personal Medicine Profile assay, genotypes obtained with the Illumina® Infinium™ Global Diversity Array platform (PMP) DNA chip were compared with in-house sequencing data for 14 pharmacogenes. Conclusions on star alleles from the results obtained by each method and the PharmVar database were identical between both technologies regarding the metabolization status, with some expected discrepancies on star allele calling: alleles *2, *9 and *36 (a *CYP2D6*::*CYP2D7* hybrid) from CYP2D6, and *4, *5, *6 and *9 alleles of *DPYD* gene (which are not tested by PMP assay). *80 allele of *UGT1A1* gene was called with PMP assay instead of *28 allele with in-house pipeline **(Table 4)**.^26^ The few variations observed are linked to limitations in the SNP-array technique (alleles tested), so this comparison is satisfactory and proves the performance of the in-house approach.

**Table 4:**
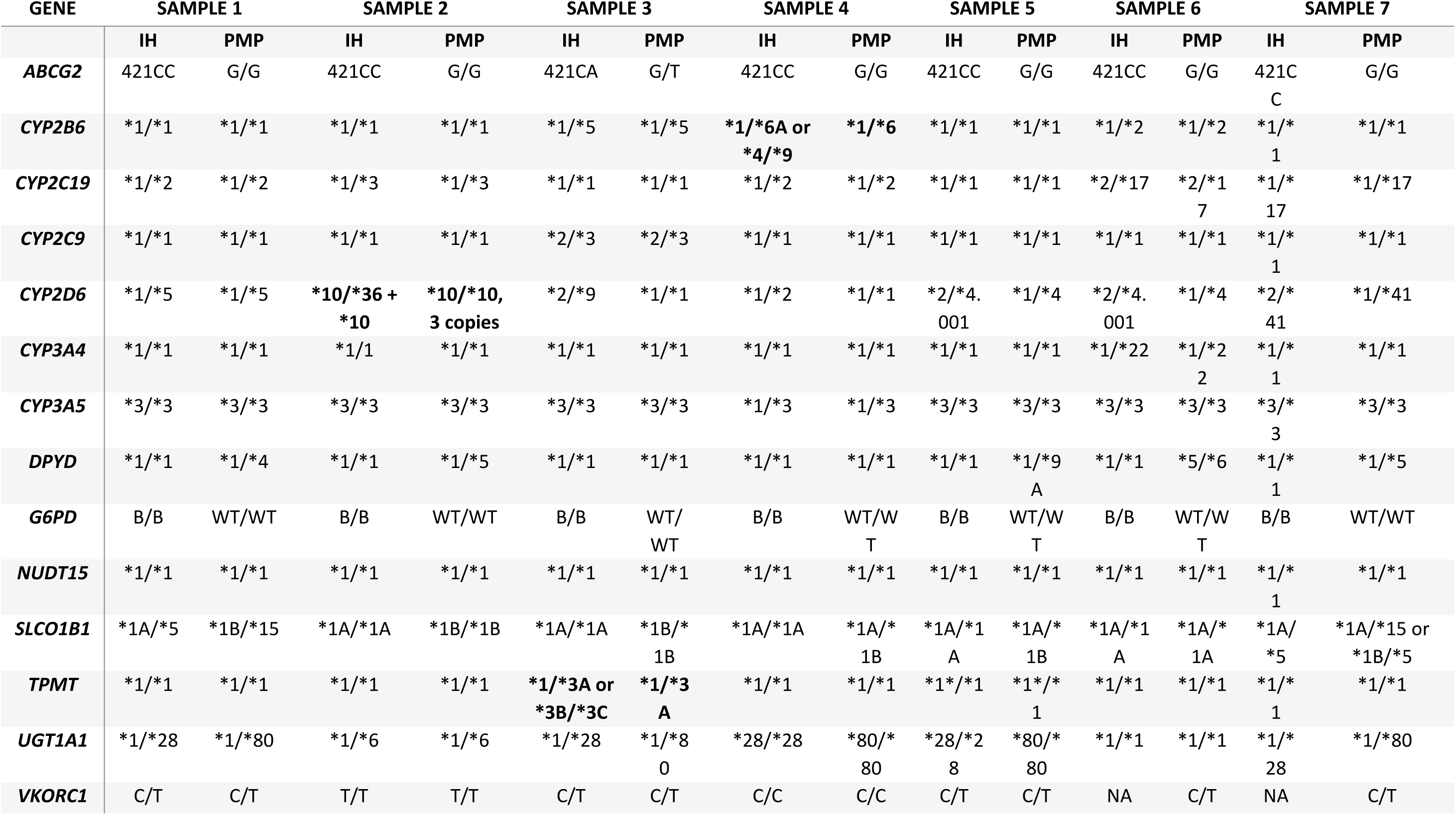
Comparison of genotyping obtained with Illumina® Infinium™ Global Diversity Array platform (PMP) and in-house (IH) flow. PMP genotyping was performed using the Illumina technology, then a report is powered by Pillcheck technology. The wildtype (mostly *1) is reported if previously listed known mutations are not detected. Analysis of 14 pharmacogenes for 7 samples are presented. Genotypes are presented with in-house workflow and compared with PMP data. Discrepancies between in-house and PMP genotype are represented in bold. *CYP2D6* *36, *2 and *9 were not tested by PMP assay, while *DPYD*\*4, *5, *6 and *9 were not tested by IH flow (associated with normal function). Moreover, *UGT1A1* *80 is tested with PMP assay as proxy for *28. *TPMT* *1/*3A and *CYP2B6* *1/*6 are an excess of interpretation from PMP (we can’t exclude *TPMT* *3B/*3C and *CYP2B6*\*4/*9 respectively, because the combination of the variants can lead to both genotypes). However, for *CYP2B6*, *4 is associated with increased function, while *6 and *9 are associated with decreased function. For *TPMT*, the patient is *1/*3A if the two variants are in the same allele, leading to an intermediate metabolizer; the patient is *3B/*3C if the variants are in two different alleles, leading to poor metabolizer. For *SLCO1B1*, *1b corresponds to *37 in the new nomenclature (normal function), *5 and *15 are both associated with absence of function. For *CYP2D6*, both *10/*36+*10 and *10/*10,3N lead to intermediate metabolizer (however, the activity score is different: 0.5 for *10/*36+*10 and 0.75 for *10/*10,3N). *36 is a hybrid CYP2D6::CYP2D7 gene without function, which is not detected by PMP.

Genotyping results obtained with PCR + LC/MS (Agena) was then compared with in-house sequencing for 33 samples, out of 48 positions analyzed by the two methods (which makes 1584 variants in total). Among the variants, 1511 results were concordant (95.4%), 66 results could not be analyzed by one of the two methods, and only 8 differences were observed (0.5%). These discrepancies are linked to the limits of Agena technique. For the *CYP2D6* gene, manual curation of sequencing reads is essential to accurately resolve the rs3892097 genotype and ensure reliable allele determination. Indeed, reads with a pattern originating from the pseudogene must be excluded (because they add wild-type background to the position). The description of the discrepancies is presented in **Table 5**. The in-house method showed consistent and reliable results.

**Table 5:**
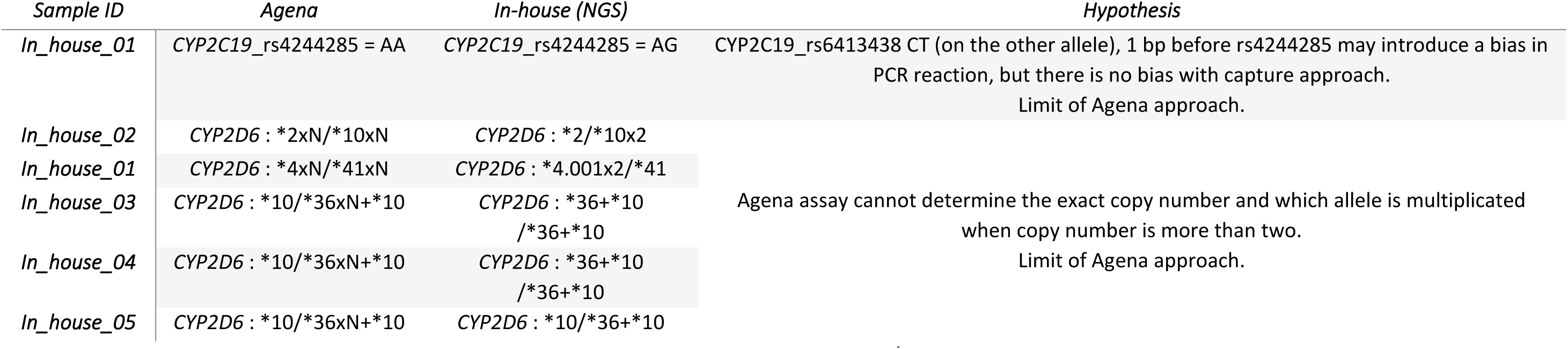
Differences observed during comparison of genotyping obtained with PCR + LC/MS (Agena) versus in-house sequencing. Among 33 samples sent to Agena for comparison, we identified 6 differences which concern 5 samples.

### *CYP2D6* star allele calling

The main analytical difficulty of the pharmacogenetic panel concerns the study of the *CYP2D6* gene due to its high similarity with the *CYP2D7* and *CYP2D8* pseudogenes. In addition, it is necessary to identify fusion genes and CNVs relating to *CYP2D6*. In order to verify the correct detection of sequence, number and structural variants in this gene, we compared star allele calling for this gene with the CDC Genetic Testing Reference Material Program (GeT-RM), a consensus of several techniques^21^. The results were compliant for all these 6 reference samples, as related in **Table 6**.

**Table 6:** Star-allele comparison of *CYP2D6* between consensus genotype and in-house (IH) genotype. For sample HG01190, the *5/*68+*4 genotype is the most likely given the frequency of association of *CYP2D6::CYP2D7* hybrid *68 with *4. Indeed, according to PharmVar database, the hybrid *68 has only been described to occur in tandem, even if we can’t exclude the existence of the *68 gene alone.

| SAMPLE | CONSENSUS<br>GENOTYPE IN GET-RM 2019 | IN-HOUSE<br>GENOTYPE | CONCLUSION |
| --- | --- | --- | --- |
| HG00421 | *2/*10x2 | *2/*10x2 | Compliant |
| HG01190 | *68+*4/*5 | *4/*68 or<br>*5/*68+*4 | Compliant<br>(same biological interpretation: poor metabolizer) |
| NA07439 | *4x2/*41 | *4.001x2<br>/*41 | Compliant |
| NA10860 | *1/*4N+*4 | *1/*4.013+*4 | Compliant<br>(*4.013 is the new nomenclature for *4N) |
| NA18545 | *5/*36x2+*10x2 | *36+*10 /*36+*10 | Compliant<br>(same biological interpretation: poor metabolizer) |
| NA12878 | *3/*68+*4 | *3/*68+*4 | Compliant |

### *UGT1A1* promotor TA-repeat calling

The number of TA repeats in the *UGT1A1* promoter was also compared. The number of TA repeats determines the level of *UGT1A1* activity (rs3064744; 7 or 8 TA repeats result in reduced activity, whereas 6 repeats result in normal function)^35^. We compared the number of TA repeats in the *UGT1A1* promoter between 5 reference sequences obtained by GS and ES in-house data.^36^ For the 5 samples, the sequencing results were consistent (**Table 7**).

**Table 7:** Comparison of TAn repeats in *UGT1A1* gene (rs3064744; TA[6]>TA[7], [8]) between reference sequences (from GeT-RM) and in-house ES data.

| SAMPLE | PMID: 36055201 | ES | CONCLUSION |
| --- | --- | --- | --- |
| NA12878 | 6/7 | 6/7 | COMPLIANT |
| HG00421 | 6/6 | 6/6 | COMPLIANT |
| HG01190 | 6/8 | 6/8 | COMPLIANT |
| NA10860 | 7/7 | 7/7 | COMPLIANT |
| NA18545 | 6/6 | 6/6 | COMPLIANT |

### HLA characterization

For the characterization of the HLA-A and HLA-B loci, a dedicated pipeline was set up (based on the xHLA tool) using exome data.^30^ The results for the NA07439 sample were compared with the genomic DNA reference materials in order to verify the conformity of the method^22^ : HLA-A*29:02:01/*68:02:01, found *29:02/*68:02 in house, and HLA-B*42:02:01/*53:01:01, found *42:02/*53:01 in house.

We carried out 2 comparisons: one with the Immucor technique already used routinely in the laboratory and accredited for HLA typing, and one with HLA results from patients who were generated by an “Etablissement Français du Sang” (EFS) laboratory in the context of a kidney transplant. When compared with the Immucor technique, the star alleles were consistent for all 14 samples tested, at the HLA-A and HLA-B loci (**Table 8**). It should be noted that a better resolution can be obtained by using a method that combines ES and xHLA than the Immucor method. xHLA is an algorithm, that improves the mapping results at the amino acid level achieving a four-digit typing accuracy for HLA genes, while the Immucor only provides a two-digit resolution.^37^ When compared with the results of the EFS laboratory for 7 transplant patients, the star alleles were identical for all the patients.

**Table 8:** Comparison of HLA-A and HLA-B characterization between ES and other techniques. a) Comparison with Immucor technique for 14 samples for which we have in-house ES data, b) comparison with results of HLA typing from EFS analysis of 7 patients with a kidney transplant

| SAMPLE | HLA-A<br>ES | - HLA-A<br>IMMUCOR | - HLA-B<br>ES | - HLA-B<br>IMMUCOR | - CONCLUSION |
| --- | --- | --- | --- | --- | --- |
| IN_HOUSE_06 | *11:01 | 11:XX | *35:01 | 35:XX | Compliant |
| IN_HOUSE_06 | *31:01 | 31:XX | *40:01 | 40:XX | Compliant |
| IN_HOUSE_07 | *30:02 | 30:XX | *15:03 | 15:XX | Compliant |
| IN_HOUSE_07 | *33:01 | 33:XX | *35:01 | 35:XX | Compliant |
| IN_HOUSE_08 | *03:01 | 03:XX | *14:02 | 14:XX | Compliant |
| IN_HOUSE_08 | *33:01 | 33:XX | *37:01 | 37:XX | Compliant |
| IN_HOUSE_09 | *02:01 | 02:XX | *14:02 | 14:XX | Compliant |
| IN_HOUSE_09 | *33:01 | 33:XX | *44:03 | 44:XX | Compliant |
| IN_HOUSE_10 | *03:01 | 03:XX | *13:01 | 13:XX | Compliant |
| IN_HOUSE_10 | *33:03 | 33:XX | *44:03 | 44:XX | Compliant |
| IN_HOUSE_11 | *23:01 | 23:XX | *53:01 | 53:XX | Compliant |
| IN_HOUSE_11 | *33:03 | 33:XX | *58:01 | 58:XX | Compliant |
| IN_HOUSE_05 | *26:01 | 26:XX | *15:02 | 15:XX | Compliant |
| IN_HOUSE_05 | *36:01 | 36:XX | *18:01 | 18:XX | Compliant |
| IN_HOUSE_12 | *31:01 | 31:XX | *40:01 | 40:XX | Compliant |
| IN_HOUSE_12 | *68:01 | 68:XX | *51:01 | 51:XX | Compliant |
| IN_HOUSE_13 | *30:02 | 30:XX | *49:01 | 49:XX | Compliant |
| IN_HOUSE_13 | *31:01 | 31:XX | *53:01 | 53:XX | Compliant |
| IN_HOUSE_14 | *01:01 | 01:XX | *40:01 | 40:XX | Compliant |
| IN_HOUSE_14 | *31:01 | 31:XX | *57:01 | 57:XX | Compliant |
| IN_HOUSE_15 | *02:01 | 02:XX | *15:02 | 15:XX | Compliant |
| IN_HOUSE_15 | *03:01 | 03:XX | *51:01 | 51:XX | Compliant |
| IN_HOUSE_16 | *01:01 | 01:XX | *08:01 | 08:XX | Compliant |
| IN_HOUSE_16 | *31:01 | 31:XX | *35:01 | 35:XX | Compliant |
| IN_HOUSE_17 | *33:01 | 33:XX | *42:02 | 42:XX | Compliant |
| IN_HOUSE_17 | *33:03 | 33:XX | *58:01 | 58:XX | Compliant |
| IN_HOUSE_18 | *23:01 | 23:XX | *44:03 | 44:XX | Compliant |
| IN_HOUSE_18 | *33:03 | 33:XX | *50:01 | 50:XX | Compliant |

**Table 8: Comparison of HLA-A and HLA-B characterization between ES and other techniques**
| SAMPLE | METHOD | HLA-A | HLA-A | HLA-B | HLA-B | CONCLUSION |
| --- | --- | --- | --- | --- | --- | --- |
| IN_HOUSE_19 | EFS | *11:01:01:01 | *66:01:01:01 | *35:01:01:02 | *41:02:01:01 | Compliant |
|  | ES | *11:01 | *66:01 | *35:01 | *41:02 | Compliant |
| IN_HOUSE_20 | EFS | *26:01:01:01 | *30:01:01:01 | *42:01:01:01 | *53:01:01:01 | Compliant |
|  | ES | *26:01 | *30:01 | *42:01 | *53:01 | Compliant |
| IN_HOUSE_21 | EFS | *03:01:01:01 | *32:01:01:01 | *40:02:01:01 | *50:02:01:01 | Compliant |
|  | ES | *03:01 | *32:01 | *40:02 | *50:02 | Compliant |
| IN_HOUSE_22 | EFS | *02:01:01 | *32:01:01 | *18:01:01 | *27:05:02 | Compliant |
|  | ES | *02:01 | *32:01 | *18:01 | *27:05 | Compliant |
| IN_HOUSE_23 | EFS | *03:01:01:07 | *30:01:01:01 | *44:03:01:01 | *58:01:01:01 | Compliant |
|  | ES | *03:01 | *30:01 | *44:03 | *58:01 | Compliant |
| IN_HOUSE_24 | EFS | *02:02:01:01 | *66:01:01:01 | *58:02:01:01 | *58:02:01:01 | Compliant |
|  | ES | *02:02 | *66:01 | *58:02 | *58:02 | Compliant |
| IN_HOUSE_25 | EFS | *11:01:01:01 | *30:01:01:01 | *35:08:01:01 | *56:01:01:03 | Compliant |
|  | ES | *11:01 | *30:01 | *35:08 | *56:01 | Compliant |

## Discussion

Pharmacogenetic analyses provide actionable information to guide drug therapy, which can be integrated with additional environmental or clinical parameters such as drug-drug interactions, renal or hepatic function and complemented by therapeutic drug monitoring (TDM) or phenotypic methods. In addition to enabling diagnosis, this type of information can be leveraged to guide therapeutic decision based on molecular analysis. Although ES is widely used in clinical settings, its application in pharmacogenetics is still limited.

Key challenges include insufficient sequencing depth and complexities linked to certain loci important for drug metabolism. Certain genes, such as *CYP2D6* and *UGT1A1*, have been seen as impossible to genotype using ES (as discussed below). Additionally, few critical pharmacogene variants are located in the intronic regions, which means they are normally not covered by ES. Despite these limitations, our various verification steps allowed us to validate the use of ES data even for regions presenting technical challenges. To our knowledge, this is the first study in which all the variants selected for their relevance in a complete pharmacogenetic panel have been analyzed solely based on ES data.

*CYP2D6* is one of the pharmacogenes that have posed significant challenges for accurate genotyping, which is particularly important considering the fact that it has a role in catabolism or bioactivation of over 25% of the clinically used drugs^7–9^. *CYP2D6* has a very high genetic variability, carrying numerous SNPs and indels that can impact gene function. However, what makes *CYP2D6* particularly hard to genotype is the presence of different structural variants (SVs), Copy Number Variants (CNVs) and hybrids between *CYP2D6* and the pseudogene *CYP2D7*. In previous studies, various bioinformatic tools have been developed for the analysis of *CYP2D6* from NGS data, but to date no study has demonstrated the possibility of studying all the haplotypes of interest in *CYP2D6* without additional analysis, particularly for the detection of hybrid genes (associated with a loss of function).^38^ Stargazer detects SNV, but also structural variations (gene deletions, duplications and conversions) from read depth information. Read depth is calculated for a control gene and for the gene of interest, enabling comparison between the two regions^39^. However, difficulties are encountered to detect *CYP2D6*::*CYP2D7* and *CYP2D7::CYP2D6* hybrid genes with this tool. This could be attributed to the limited reliability of using another gene from the same sample as an internal control. In our method, control depth is calculated within a large cohort of 1246 samples. StellarPGx is able to detect variants from 12 pharmacogenes including *CYP2D6*, however false positive and false negative results were observed for genotypes containing hybrid genes^40^. In our study, we were able to detect both SNV and SV, including SNV and hybrid genes. When comparing the *CYP2D6* genotypes with PMP assay, discrepancies were observed due to missing allele detection (*2, *9, hybrid *36) with the PMP assay. When comparing with PCR + LC/MS approach (Agena), the only discrepancies were due to the limitation of the Agena approach, which cannot determine the exact copy number and which allele is multiplicated when the copy number exceeds two. *UGT1A1* genotype is an important determinant of irinotecan toxicity and guide dose adjustment in clinical practice. Specifically, the *6 and *28 alleles are strongly associated with incidence of adverse events related with irinotecan^41^. The *28 Indel variant is complex to analyze due to a potential lack of coverage with ES in this region^5^. In our study, we overcame this difficulty by manually checking for the presence of indel in *UGT1A1* by viewing the BAM files in IGV. No discrepancies were found when comparing with other methods. However, for the moment our method does not allow us to analyze without a manual verification step for this region.

One of the previously described challenges when it comes to the use of ES to obtain a pharmacogenetic profile is low sequencing depth compared with targeted analysis. We addressed this by analyzing the pharmacogenetic profile as part of the diagnostic process, using a technique that is highly reproducible and can be performed on a large number of samples. The depth is therefore not only homogenous, but also substantial (150X) on average. Another factor that may explain the success of pharmacogenetic analysis using our method is the bioinformatics pipeline, which was optimized as we progressed (using GATK4). Finally, manual curation downstream enables us to resolve any remaining ambiguities linked to loci that are difficult to interpret (*CYP2D6 and UGT1A1*).

This method focuses on a predefined list of variants with established phenotype associations. This panel is characterized by a high diversity of detected variants, making it suitable as a proof of concept. However, because it is not targeted sequencing, additional variants may be detected, raising questions regarding the interpretation of these less well-characterized findings. The contribution of NGS for the detection and description of novel variants in pharmacogenes has already been demonstrated, particularly for the loss of function variants^42^. For novel missense variants, we can rely on bioinformatical prediction tools (based on frequency in control population, expected impact on the protein conformation, potential impact on splicing), but we need more information to provide a proper interpretation. Once published, new variants can be added to the list without having to perform a new technique. The list of variants in the pharmacogenetic panel is not fixed, allowing new variants to be added and the panel to be adapted as needed.

It is important to note that the pharmacogenetic profile search is intended to be carried out on the basis of a specific request from the prescriber. It requires clear information to be given to the patient beforehand and written consent to be obtained. This information cannot be returned as secondary data.

## Conclusion

Our study demonstrates that exome sequencing can be leveraged far beyond its traditional role in genetic diagnosis, providing at the same time a complete, reliable, and comprehensive characterization of the patient’s pharmacogenomic profile. By overcoming long-standing barriers such as low sequencing depth and the complexity of *CYP2D6*, *UGT1A1*, and HLA loci, we show that a single test can deliver all the information required both for etiological investigation and for guiding therapeutic decisions.

This approach positions exome sequencing as a true cornerstone of precision medicine: it enables the anticipation of drug response, retrospective interpretation of pharmacogenomic phenotypes, and paves the way for systematic integration of pharmacogenomics into clinical practice. By unlocking the full value of NGS data, it offers an unprecedented opportunity to unify molecular diagnosis and pharmacogenomics into a single, standardized workflow that can directly benefit patient care.

It is essential for this information to be complemented by phenotypic methods which can be recommended or even mandatory in specific clinical context and to be considered alongside therapeutic guidelines to ensure appropriate drug use. Furthermore, it should be integrated with other critical data (such as renal or hepatic function and drug-drug interactions) to specifically meet the needs of diverse patient profiles.

## Ethics declaration

The study was approved by an Institutional Review Board (Direction de la Recherche Clinique et de l’Innovation (APHP220461)) and the Ethics board of Sorbonne Université (CER-2022-009).

Written informed consent for participation in the study and publication of related data was obtained from all individuals whose data are included in the manuscript, in accordance with institutional, national, and international regulations.

Copies of all consent forms are securely stored and can be made available to the journal or relevant authorities upon request.

## Data sharing statement

Anonymized data that support the findings of this study are available on request from the corresponding author, Laure Raymond. The data are not publicly available due to restrictions, their containing information that could compromise the privacy of research participants.

## References

1. Manson LE, Van Der Wouden CH, Swen JJ, Guchelaar HJ. The Ubiquitous Pharmacogenomics consortium: making effective treatment optimization accessible to every European citizen. Pharmacogenomics. 2017;18(11):1041–1045. doi:10.2217/pgs-2017-0093

2. Iemmolo R, La Cognata V, Morello G, et al. Development of a Pharmacogenetic Lab-on-Chip Assay Based on the In-Check Technology to Screen for Genetic Variations Associated to Adverse Drug Reactions to Common Chemotherapeutic Agents. Biosensors. 2020;10(12):202.doi:10.3390/bios10120202

3. Williams GR, Cook L, Lewis LD, Tsongalis GJ, Nerenz RD. Clinical Validation of a 106-SNV MALDI-ToF MS Pharmacogenomic Panel. The Journal of Applied Laboratory Medicine. 2020;5(3):454–466. doi:10.1093/jalm/jfaa018

4. Londin ER, Clark P, Sponziello M, Kricka LJ, Fortina P, Park JY. Performance of exome sequencing for pharmacogenomics. Personalized Medicine. 2015;12(2):109–115. doi:10.2217/pme.14.77

5. Lanillos J, Carcajona M, Maietta P, Alvarez S, Rodriguez-Antona C. Clinical pharmacogenetic analysis in 5,001 individuals with diagnostic Exome Sequencing data. npj Genom Med. 2022;7(1):12. doi:10.1038/s41525-022-00283-3

6. Thauvin-Robinet C, Thevenon J, Nambot S, et al. Secondary actionable findings identified by exome sequencing: expected impact on the organisation of care from the study of 700 consecutive tests. Eur J Hum Genet. 2019;27(8):1197–1214. doi:10.1038/s41431-019-0384-7

7. Turner AJ, Nofziger C, Ramey BE, et al. PharmVar Tutorial on *CYP2D6* Structural Variation Testing and Recommendations on Reporting. Clin Pharma and Therapeutics. 2023;114(6):1220–1237. doi:10.1002/cpt.3044

8. Yang Y, Botton MR, Scott ER, Scott SA. Sequencing the *CYP2D6* gene: from variant allele discovery to clinical pharmacogenetic testing. Pharmacogenomics. 2017;18(7):673–685. doi:10.2217/pgs-2017-0033

9. Rakela J, Rule J, Ganger D, et al. Whole Exome Sequencing Among 26 Patients With Indeterminate Acute Liver Failure: A Pilot Study. Clin Transl Gastroenterol. 2019;10(10):e00087. doi:10.14309/ctg.0000000000000087

10. Stüven T, Griese EUi, Kroemer HK, Eichelbaum M, Zanger UM. Rapid detection of CYP2D6 null alleles by long distance-and multiplex-polymerase chain reaction: Pharmacogenetics. 1996;6(5):417–421. doi:10.1097/00008571-199610000-00005

11. Van Der Lee M, Allard WG, Bollen S, et al. Repurposing of Diagnostic Whole Exome Sequencing Data of 1,583 Individuals for Clinical Pharmacogenetics. Clin Pharma and Therapeutics. 2020;107(3):617–627. doi:10.1002/cpt.1665

12. Butler-Laporte G, Farjoun J, Nakanishi T, et al. HLA allele-calling using multi-ancestry whole-exome sequencing from the UK Biobank identifies 129 novel associations in 11 autoimmune diseases. Commun Biol. 2023;6(1):1113. doi:10.1038/s42003-023-05496-5

13. Abdullah-Koolmees H, Van Keulen AM, Nijenhuis M, Deneer VHM. Pharmacogenetics Guidelines: Overview and Comparison of the DPWG, CPIC, CPNDS, and RNPGx Guidelines. Front Pharmacol. 2021;11:595219. doi:10.3389/fphar.2020.595219

14. Pratt VM, Del Tredici AL, Hachad H, et al. Recommendations for Clinical CYP2C19 Genotyping Allele Selection: A Report of the Association for Molecular Pathology. J Mol Diagn. 2018;20(3):269–276. doi:10.1016/j.jmoldx.2018.01.011

15. Pratt VM, Cavallari LH, Del Tredici AL, et al. Recommendations for Clinical Warfarin Genotyping Allele Selection: A Report of the Association for Molecular Pathology and the College of American Pathologists. J Mol Diagn. 2020;22(7):847–859. doi:10.1016/j.jmoldx.2020.04.204

16. Pratt VM, Cavallari LH, Del Tredici AL, et al. Recommendations for Clinical CYP2C9 Genotyping Allele Selection: A Joint Recommendation of the Association for Molecular Pathology and College of American Pathologists. J Mol Diagn. 2019;21(5):746–755. doi:10.1016/j.jmoldx.2019.04.003

17. Pratt VM, Cavallari LH, Del Tredici AL, et al. Recommendations for Clinical CYP2D6 Genotyping Allele Selection: A Joint Consensus Recommendation of the Association for Molecular Pathology, College of American Pathologists, Dutch Pharmacogenetics Working Group of the Royal Dutch Pharmacists Association, and the European Society for Pharmacogenomics and Personalized Therapy. J Mol Diagn. 2021;23(9):1047–1064. doi:10.1016/j.jmoldx.2021.05.013

18. Pratt VM, Cavallari LH, Fulmer ML, et al. CYP3A4 and CYP3A5 Genotyping Recommendations: A Joint Consensus Recommendation of the Association for Molecular Pathology, Clinical Pharmacogenetics Implementation Consortium, College of American Pathologists, Dutch Pharmacogenetics Working Group of the Royal Dutch Pharmacists Association, European Society for Pharmacogenomics and Personalized Therapy, and Pharmacogenomics Knowledgebase. J Mol Diagn. 2023;25(9):619–629. doi:10.1016/j.jmoldx.2023.06.008

19. Pratt VM, Cavallari LH, Fulmer ML, et al. DPYD Genotyping Recommendations: A Joint Consensus Recommendation of the Association for Molecular Pathology, American College of Medical Genetics and Genomics, Clinical Pharmacogenetics Implementation Consortium, College of American Pathologists, Dutch Pharmacogenetics Working Group of the Royal Dutch Pharmacists Association, European Society for Pharmacogenomics and Personalized Therapy, Pharmacogenomics Knowledgebase, and Pharmacogene Variation Consortium. J Mol Diagn. 2024;26(10):851–863. doi:10.1016/j.jmoldx.2024.05.015

20. Pratt VM, Cavallari LH, Fulmer ML, et al. TPMT and NUDT15 Genotyping Recommendations: A Joint Consensus Recommendation of the Association for Molecular Pathology, Clinical Pharmacogenetics Implementation Consortium, College of American Pathologists, Dutch Pharmacogenetics Working Group of the Royal Dutch Pharmacists Association, European Society for Pharmacogenomics and Personalized Therapy, and Pharmacogenomics Knowledgebase. J Mol Diagn. 2022;24(10):1051–1063. doi:10.1016/j.jmoldx.2022.06.007

21. Gaedigk A, Turner A, Everts RE, et al. Characterization of Reference Materials for Genetic Testing of CYP2D6 Alleles. The Journal of Molecular Diagnostics. 2019;21(6):1034–1052. doi:10.1016/j.jmoldx.2019.06.007

22. Bettinotti MP, Ferriola D, Duke JL, et al. Characterization of 108 Genomic DNA Reference Materials for 11 Human Leukocyte Antigen Loci. The Journal of Molecular Diagnostics. 2018;20(5):703–715. doi:10.1016/j.jmoldx.2018.05.009

23. Eberle MA, Fritzilas E, Krusche P, et al. A reference data set of 5.4 million phased human variants validated by genetic inheritance from sequencing a three-generation 17-member pedigree. Genome Res. 2017;27(1):157–164. doi:10.1101/gr.210500.116

24. Pratt VM, Everts RE, Aggarwal P, et al. Characterization of 137 Genomic DNA Reference Materials for 28 Pharmacogenetic Genes. The Journal of Molecular Diagnostics. 2016;18(1):109–123. doi:10.1016/j.jmoldx.2015.08.005

25. Lois A, Everts R, Nakorchevsky A, Aleksey A, Hunt P, Jackson K. The VeriDose Core Panel: Strong Performance When Analyzing 438 Challenging Pharmacogenetic Samples. Published online April 2, 2021. https://agenabio.com/wp-content/uploads/2020/06/Agena-Conference-Poster-Veridose-Core-V3-G020.pdf

26. Bravo-Gómez A, Salvador-Martín S, Zapata-Cobo P, Sanjurjo-Sáez M, López-Fernández LA. Genotyping of UGT1A1*80 as an Alternative to UGT1A1*28 Genotyping in Spain. Pharmaceutics. 2022;14(10):2082. doi:10.3390/pharmaceutics14102082

27. Yaldiz B, Kucuk E, Hampstead J, et al. Twist exome capture allows for lower average sequence coverage in clinical exome sequencing. Hum Genomics. 2023;17(1):39. doi:10.1186/s40246-023-00485-5

28. Testard Q, Vanhoye X, Yauy K, et al. Exome sequencing as a first-tier test for copy number variant detection: retrospective evaluation and prospective screening in 2418 cases. J Med Genet. 2022;59(12):1234–1240. doi:10.1136/jmg-2022-108439

29. DePristo MA, Banks E, Poplin R, et al. A framework for variation discovery and genotyping using next-generation DNA sequencing data. Nat Genet. 2011;43(5):491–498. doi:10.1038/ng.806

30. Xie C, Yeo ZX, Wong M, et al. Fast and accurate HLA typing from short-read next-generation sequence data with xHLA. Proc Natl Acad Sci U S A. 2017;114(30):8059–8064. doi:10.1073/pnas.1707945114

31. Robinson JT, Thorvaldsdottir H, Turner D, Mesirov JP. igv.js: an embeddable JavaScript implementation of the Integrative Genomics Viewer (IGV). Alkan C, ed. Bioinformatics. 2023;39(1):btac830. doi:10.1093/bioinformatics/btac830

32. Gel B, Serra E. karyoploteR: an R/Bioconductor package to plot customizable genomes displaying arbitrary data. Bioinformatics. 2017;33(19):3088–3090. doi:10.1093/bioinformatics/btx346

33. Rehm HL, Bale SJ, Bayrak-Toydemir P, et al. ACMG clinical laboratory standards for next-generation sequencing. Genetics in Medicine. 2013;15(9):733–747. doi:10.1038/gim.2013.92

34. Eberle MA, Fritzilas E, Krusche P, et al. A reference data set of 5.4 million phased human variants validated by genetic inheritance from sequencing a three-generation 17-member pedigree. Genome Res. 2017;27(1):157–164. doi:10.1101/gr.210500.116

35. Vukovic M, Radlovic N, Lekovic Z, et al. *UGT1A1* (TA) _n_ promoter genotype: Diagnostic and population pharmacogenetic marker in Serbia. Balkan Journal of Medical Genetics. 2018;21(1):59–68. doi:10.2478/bjmg-2018-0012

36. Byrska-Bishop M, Evani US, Zhao X, et al. High-coverage whole-genome sequencing of the expanded 1000 Genomes Project cohort including 602 trios. Cell. 2022;185(18):3426–3440.e19. doi:10.1016/j.cell.2022.08.004

37. Xie C, Yeo ZX, Wong M, et al. Fast and accurate HLA typing from short-read next-generation sequence data with xHLA. Proc Natl Acad Sci USA. 2017;114(30):8059–8064. doi:10.1073/pnas.1707945114

38. Hari A, Zhou Q, Gonzaludo N, et al. An efficient genotyper and star-allele caller for pharmacogenomics. Genome Res. 2023;33(1):61–70. doi:10.1101/gr.277075.122

39. Lee S been, Wheeler MM, Patterson K, et al. Stargazer: a software tool for calling star alleles from next-generation sequencing data using CYP2D6 as a model. Genetics in Medicine. 2019;21(2):361–372. doi:10.1038/s41436-018-0054-0

40. Twesigomwe D, Drögemöller BI, Wright GEB, et al. StellarPGx: A Nextflow Pipeline for Calling Star Alleles in Cytochrome P450 Genes. Clin Pharma and Therapeutics. 2021;110(3):741–749. doi:10.1002/cpt.2173

41. Emery LP, Brooks GA. Revisiting *UGT1A1* Pharmacogenetic Testing Before Irinotecan—Why Not? JCO Oncology Practice. 2022;18(4):281–282. doi:10.1200/OP.21.00840

42. Ingelman-Sundberg M, Mkrtchian S, Zhou Y, Lauschke VM. Integrating rare genetic variants into pharmacogenetic drug response predictions. Hum Genomics. 2018;12(1):26. doi:10.1186/s40246-018-0157-3

